# Divergent Cognitive Trajectories by Gamma Center Frequency Plasticity After Personalized Gamma Entrainment in Early Alzheimer Disease: A Dechallenge Analysis

**DOI:** 10.64898/2026.04.28.26351905

**Authors:** Yeseung Park, Ji Won Han, Myeonghae Hyun, Ki Woong Kim

## Abstract

**Background:** Non-invasive gamma entrainment using sensory stimulation (GENUS) is being investigated as a therapy for Alzheimer disease (AD), but the clinical course of participants who fail to show gamma center frequency plasticity remains unclear. We therefore examined whether cognitive decline observed during personalized GENUS was attenuated after cessation in participants stratified post hoc by CF change.

**Methods:** This case series was derived from an open-label proof-of-concept trial with extended follow-up (mean 26.3 months). Sixteen participants with amyloid-positive early AD completed 12 weeks of home-based daily flickering light stimulation (1 hour/day) at individualized gamma frequency, and 12 completed long-term follow-up. Participants were classified post hoc as ICF+ (CF increase ≥2 Hz; n=5) or ICF− (no CF increase; n=7). MMSE trajectories from baseline to week 12 and from week 12 to final follow-up were analyzed using piecewise linear mixed-effects models.

**Results:** Baseline characteristics, including MMSE, did not differ significantly between groups. During intervention, MMSE was stable in ICF+ (+0.27 points/month; 95% CI, −0.20 to 0.73) but declined in ICF− (−0.69 points/month; 95% CI, −1.07 to −0.30; between-group p=0.011). After cessation, estimated slopes were similar in ICF+ and ICF− (−0.16 vs −0.17 points/month; p=0.968), and the phase×group interaction was significant (p=0.006). Medication intensification during follow-up was more common in ICF−, including antipsychotic initiation in 4 of 7 participants.

**Conclusions:** In this exploratory post hoc analysis, lack of CF plasticity was associated with accelerated cognitive decline during the intervention phase but not during follow-up. This temporal pattern is consistent with, but does not establish, a dechallenge-like safety signal. Given the small sample, post hoc stratification, and differential medication changes, these findings should be considered hypothesis-generating and require prospective validation with pre-defined electrophysiologic stratification.

**Trial registration:** Clinical Research Information Service (CRIS), Republic of Korea (KCT0010618); submitted 6 October 2022; first patient enrolled 2 February 2023. https://cris.nih.go.kr/cris/search/detailSearch.do?seq=31321.

## Introduction

Non-invasive gamma entrainment using sensory stimulation (GENUS) is being investigated as a potential therapy for Alzheimer disease (AD)^1^, but the clinical significance of heterogeneous electrophysiologic responses remains uncertain. In our pilot study of personalized GENUS, all participants demonstrated acute gamma entrainment at baseline but exhibited divergent trajectories after the 12-week intervention^2^. In our prior pilot study, participants showed divergent post-intervention changes in endogenous gamma center frequency (CF), and those without CF increase appeared to experience greater cognitive decline during the intervention period^2^. These findings raise the possibility that failure to show CF plasticity may be associated with a different clinical course during gamma entrainment, rather than representing a neutral absence of benefit. If so, the absence of electrophysiological response may constitute a marker of vulnerability rather than a neutral outcome^3^. Here, we report extended follow-up data to examine whether the decline observed during the intervention period was attenuated after cessation. We treated this as an exploratory, hypothesis-generating dechallenge analysis rather than a causal test of harm.

## Methods

### Study Design and Participants

This study reports long-term safety outcomes of an open-label, proof-of-concept trial of personalized GENUS (CRIS No. KCT0010618). Enrollment occurred from February 6 to December 15, 2023. The study was approved by the Institutional Review Board of Seoul National University Bundang Hospital (IRB No. B-2302-810-302). Written informed consent was obtained from all participants. Sixteen individuals with amyloid-positive early AD completed a 12-week home-based GENUS intervention. Individuals without acute entrainment at baseline (signal-to-noise ratio < 1.5) were excluded. Of the 16 completers, 12 were available for long-term follow-up (mean 26.3 ± 4.5 months).

Participants self-administered flickering light stimulation for 1 hour daily at their individualized gamma frequency (32–40 Hz). For the present follow-up analysis, participants were classified post hoc according to gamma CF change from baseline to week 12: ICF+ was defined as a CF increase of ≥2 Hz, and ICF− as no CF increase despite confirmed entrainment capacity and adherence. Because this classification was based on a post-intervention electrophysiologic outcome, all between-group comparisons should be interpreted as exploratory^2^.

### Statistical Analyses

Baseline characteristics were compared using Mann-Whitney U tests and Fisher exact tests. Given the pilot nature of the study, no formal sample size calculation was performed and all analyses were considered hypothesis-generating. Cognitive trajectories were analyzed using piecewise linear mixed-effects models (PLMM)^4^ with two phases: intervention (Phase 1; baseline to week 12) and post-intervention (Phase 2; week 12 to final assessment). The model included random intercepts and Phase 2 random slopes for inter-individual variability in decline trajectories^5^. Fixed effects included time, group, time-by-group interactions, and covariates (age, sex, and education). The primary comparison was whether the difference in MMSE slopes between ICF+ and ICF− varied across the two phases. Given the small sample size and the post hoc nature of subgrouping, these analyses were intended to describe temporal patterns rather than to establish causality. Analyses were performed using R (version 4.3.2) with statistical significance set at two-tailed p < 0.05.

### Data Availability

Data are not publicly available, but access may be considered upon reasonable request and subject to institutional approval.

### Standard Protocol Approvals, Registrations, and Patient Consents

Ethics approval: This study was approved by the Institutional Review Board of Seoul National University Bundang Hospital (IRB no. B-2302-810-302).

Informed consent: Written informed consent for research participation was obtained from all participants (or their legally authorized representatives, when applicable).

Trial registration: The trial was registered with the Clinical Research Information Service (CRIS) (KCT0010618).

Enrollment period: Enrollment occurred from February 2, 2023 to December 15, 2023.

## Results

Twelve participants (5 ICF+, 7 ICF−) completed long-term follow-up (26.3 ± 4.5 months). Baseline characteristics did not differ significantly between groups (**Table 1**). However, because subgroup classification was defined post hoc and the sample was small, the absence of statistically significant baseline differences should not be interpreted as proof of equivalence between groups. Medication trajectories also differed during follow-up and should be considered when interpreting post-intervention MMSE trends. While baseline donepezil dosages were comparable (p=0.848), the ICF– group required greater escalations (+2.9 mg vs. 0.0 mg; p=0.318), with all ICF+ participants maintaining stable dosages. Psychotropic burden showed a similar pattern: the ICF+ group had three discontinuations against one increase, whereas the ICF– group required ten changes (five additions, five escalations) with none. Antipsychotic initiation (quetiapine/aripiprazole) occurred in 57.1% of ICF– participants (n=4/7) for emergent behavioral and psychological symptoms of dementia; no ICF+ participants required antipsychotics. The ICF+ group achieved a net reduction in benzodiazepine use, while all ICF– users required maintenance or escalation.

**Table 1.**
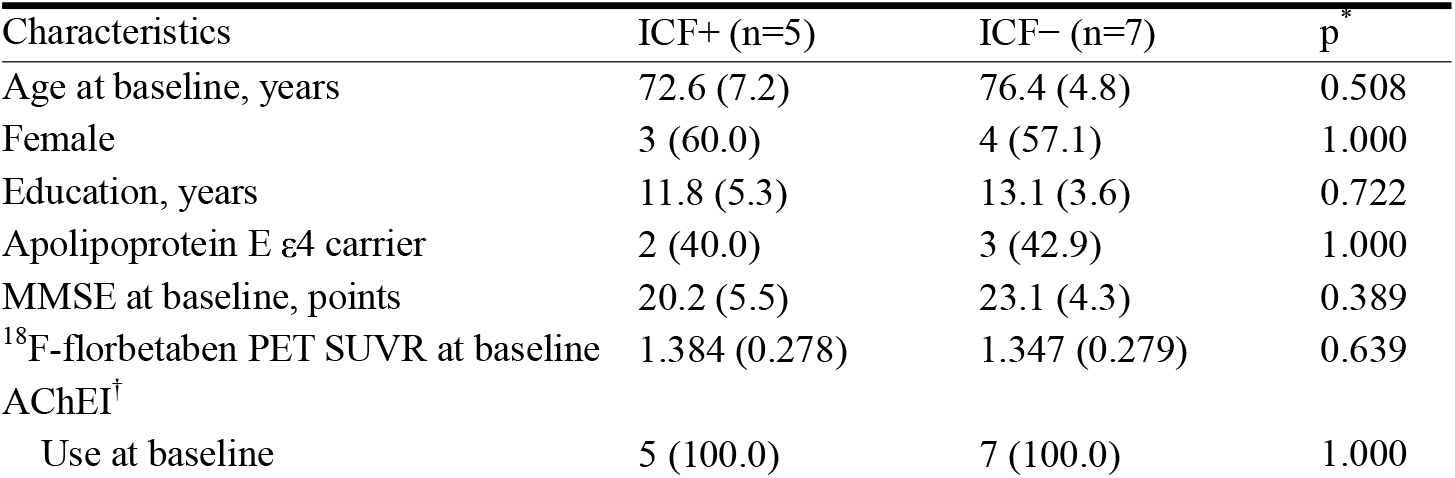
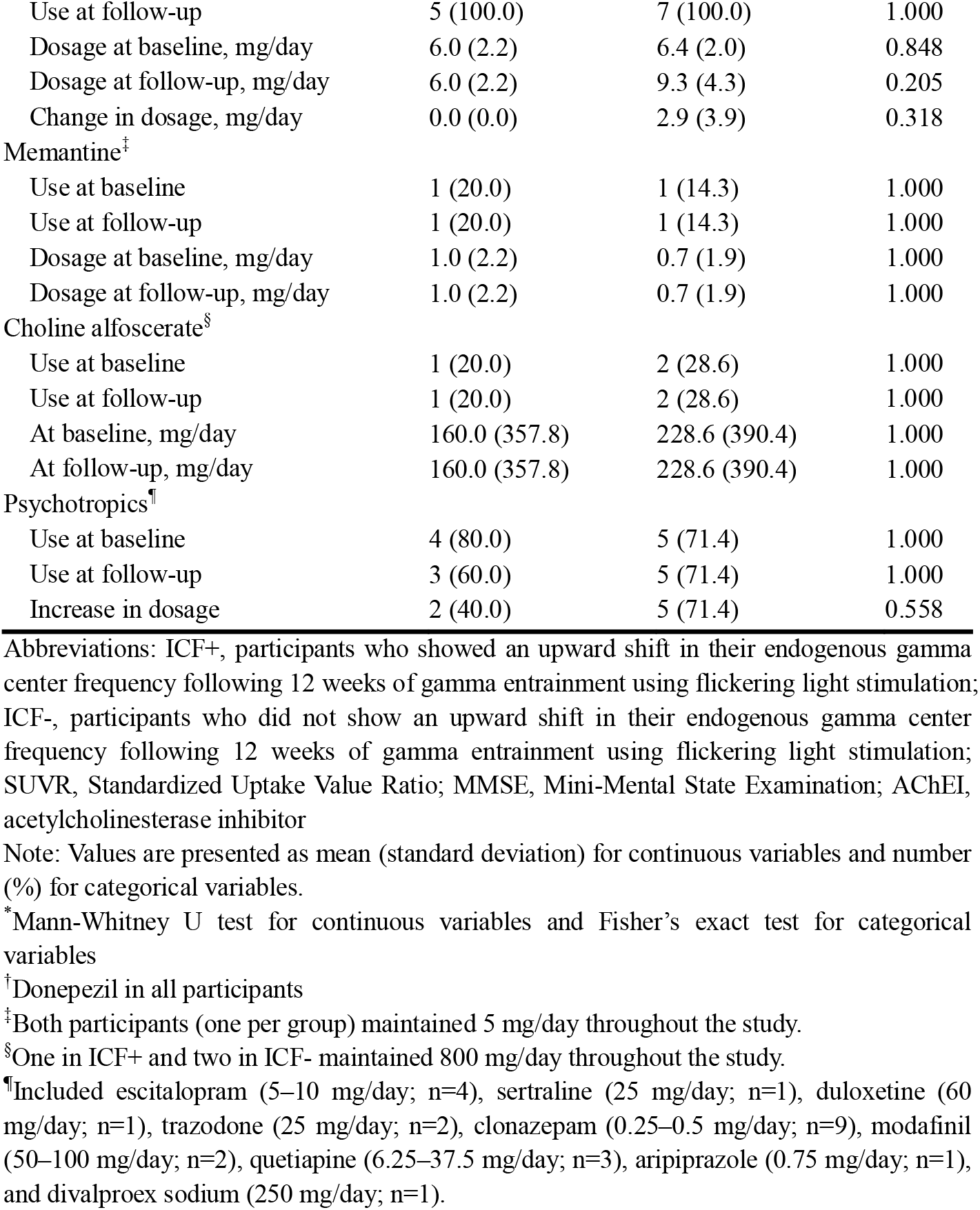
Characteristics of Study Participants.

PLMM revealed divergent cognitive trajectories during the intervention phase (**Figure 1**). The ICF+ group showed no significant change (slope = +0.27 points/month; 95% CI, −0.20 to 0.73), consistent with cognitive stability, while the ICF− group exhibited significant decline (slope = −0.69 points/month; 95% CI, −1.07 to −0.30; between-group p=0.011). The time×group interaction was significant (p=0.006), indicating greater divergence during intervention than post-intervention follow-up. The estimated net difference between groups over the 12-week intervention was approximately 2.7 MMSE points. Given the small sample size and post hoc subgrouping, this finding should be interpreted as a preliminary between-group difference rather than definitive evidence of harm.

**Figure 1.**
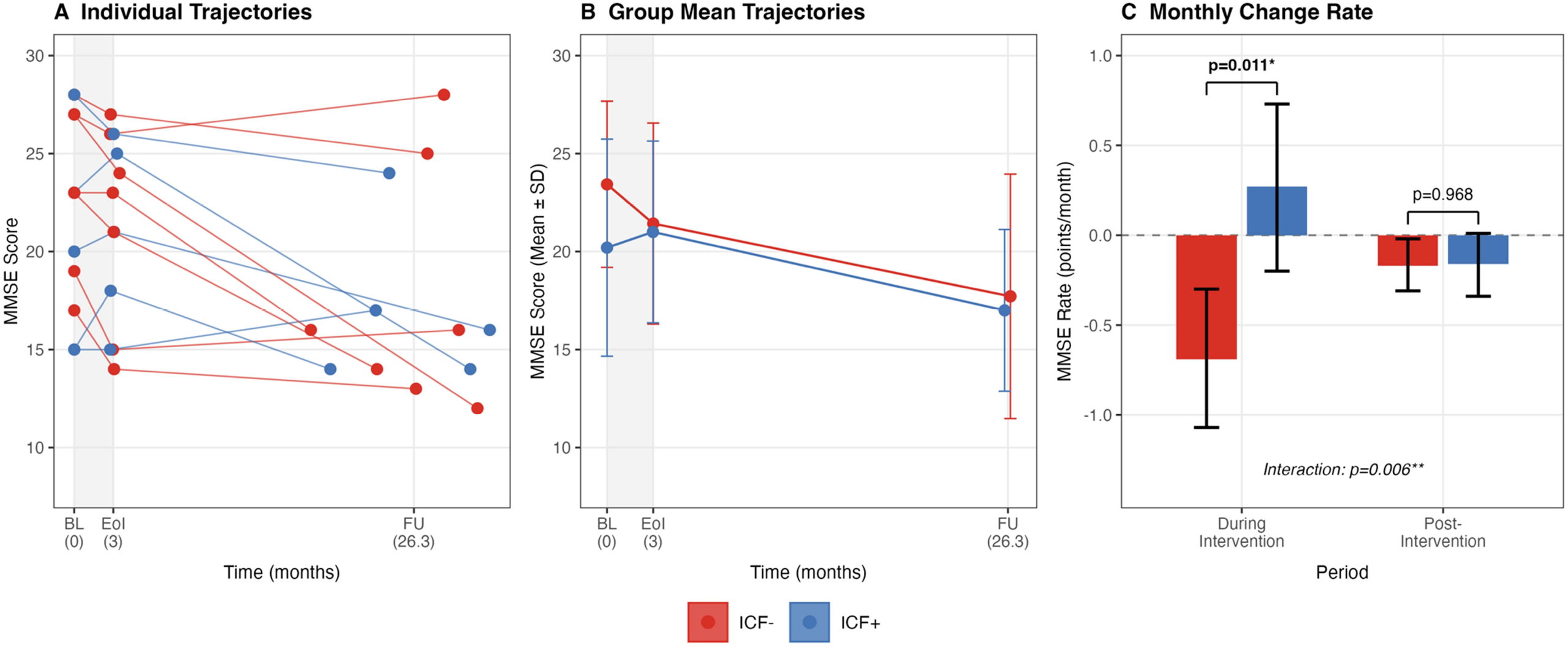
Cognitive Trajectories Following Gamma Entrainment Therapy Stratified by Electrophysiological Response. MMSE scores were tracked from baseline through the end of intervention (∼3 months) to long-term follow-up (∼26 months). Participants were stratified into ICF+ (responders, n=5) and ICF-(non-responders, n=7) groups based on whether they exhibited ≥2 Hz increase in endogenous gamma center frequency following the intervention. (A) Individual MMSE trajectories for all participants. Each line represents one participant’s cognitive course over time. (B) Group mean MMSE trajectories with error bars representing standard deviation. (C) Monthly MMSE change rates estimated from piecewise linear mixed-effects models with random intercepts and random slopes for the post-intervention phase. Error bars represent 95% confidence intervals. Shaded areas in panels A and B indicate the intervention period. Abbreviations: BL, baseline; EoI, end of intervention; FU, follow-up; ICF, increase in center frequency after gamma entrainment intervention; MMSE, Mini-Mental State Examination.

The accelerated deterioration in the ICF− group was temporally confined to the intervention period. Upon cessation, both groups showed similar decline rates: ICF+ at −0.16 points/month (95% CI, −0.34 to 0.01) and ICF− at −0.17 points/month (95% CI, −0.31 to −0.02; between-group p = 0.968). This convergence is consistent with a dechallenge-like temporal pattern, but it may also reflect medication-related stabilization or other time-varying clinical factors rather than a pure effect of stimulation cessation. Despite the similarity of estimated post-intervention slopes, the MMSE difference accrued during the intervention period was not recovered during follow-up. This observation should be interpreted cautiously because the study was exploratory and medication changes were not balanced between groups.

Although baseline MMSE did not differ significantly between groups (p=0.389), higher baseline MMSE in ICF− (23.1 vs 20.2 points) should be considered when interpreting change scores in this small sample. The PLMM accounted for baseline differences through random intercepts, but residual interpretive uncertainty remains.

## Discussion

In this extended follow-up of an open-label personalized GENUS trial, participants who did not show an increase in gamma CF exhibited faster MMSE decline during the intervention phase, whereas slope differences were not evident during follow-up. These findings suggest electrophysiologic heterogeneity in clinical course during exposure to personalized gamma entrainment, but do not establish definitive harm^6^.

The dechallenge pattern provides the most compelling evidence that this acceleration was temporally associated with the intervention period. If ICF− participants were intrinsic rapid progressors, their precipitous decline (−0.69 points/month; p=0.011) should have persisted after cessation. Instead, their trajectory flattened to −0.17 points/month upon discontinuation, implicating active stimulation as the driver of transient deterioration.

Alternative explanations remain plausible. Regression to the mean, natural disease variability, and medication intensification in the ICF− group, including antipsychotic initiation in 4 of 7 participants and greater donepezil escalation, may all have contributed to the apparent convergence of post-intervention slopes. Because assessments were not timed to distinguish an immediate dechallenge effect from delayed pharmacologic stabilization, the current design cannot disentangle these possibilities. Comparable baseline amyloid burden and APOE ε4 status nonetheless argue against inherently more aggressive pathology in the ICF− group.

The similarity of post-intervention slopes does not negate the observation that cognitive loss accrued during the intervention period was not recovered during the follow-up window. Rather than indicating definitive harm, this pattern suggests that lack of electrophysiologic plasticity may identify a subgroup warranting closer monitoring in future prospective studies.

We hypothesize that this effect may stem from metabolic excitotoxicity. Gamma oscillations demand high ATP consumption for inhibitory-excitatory coupling^7,8^, and in AD brains with compromised neurovascular coupling^9^ and mitochondrial dysfunction^10^, forced high-frequency synchronization may create a critical mismatch between metabolic demand and supply, precipitating synaptic fatigue and oxidative stress^11^. If replicated prospectively, baseline electrophysiologic profiling may have value not only for treatment stratification but also for safety monitoring during personalized gamma entrainment.

### Limitations

Several limitations merit consideration. First, the sample was small (n=12; ICF+ n=5, ICF− n=7), limiting precision and increasing the fragility of interaction and effect-size estimates. The proposed metabolic excitotoxicity mechanism remains hypothetical without direct metabolic measurements. Alternative explanations including intervention-related psychological stress, circadian disruption, or nocebo effects cannot be excluded. Second, ICF classification was performed post hoc using change in gamma CF from baseline to week 12, which introduces circularity and may have inflated the observed phase×group interaction. Accordingly, we cannot determine whether ICF status represents differential response to stimulation or a proxy for unmeasured disease characteristics. Third, medication changes in the ICF− group, particularly greater donepezil escalation and antipsychotic initiation, represent important time-varying confounders. These adjustments may have contributed to the apparent post-intervention stabilization and limit causal interpretation of a dechallenge-like pattern. Fourth, the study was open-label and lacked a sham or untreated comparator, so the observed trajectories cannot be cleanly separated from regression to the mean, natural disease progression, or other non-specific influences. Fifth, MMSE was the only cognitive outcome analyzed in this report and may be influenced by behavioral or symptomatic factors. Taken together, these limitations indicate that the present findings should be interpreted as preliminary and hypothesis-generating rather than as definitive evidence of harm.

## Conclusions

In this exploratory post hoc follow-up analysis, lack of gamma center frequency plasticity was associated with accelerated MMSE decline during the intervention phase but not during follow-up. These findings are consistent with a hypothesis-generating safety signal and warrant prospective validation using pre-defined electrophysiologic stratification and careful monitoring of concurrent medication changes.

## Data Availability

All data produced in the present study are available upon reasonable request to the authors

## Funding

This work was supported by the Engineering Research Center of Excellence (ERC) Program through the National Research Foundation of Korea (NRF), funded by the Ministry of Science and ICT (MSIT) (Grant No. NRF-2017R1A5A1014708), and by a grant of the Korea Health Technology R&D Project through the Korea Health Industry Development Institute (KHIDI), funded by the Ministry of Health & Welfare, Republic of Korea (Grant No. RS-2025-02223212). The sponsors had no role in the design and conduct of the study; in the collection, analysis, and interpretation of data; in the preparation of the manuscript; or in the review or approval of the manuscript.

### Declaration of Generative AI and AI-assisted technologies in the writing process

During the preparation of this manuscript, the authors used ChatGPT (OpenAI) for English-language editing and refinement of sentence structure. The use of this AI-assisted tool was limited to the writing process and accounted for approximately 10% of manuscript preparation. The authors reviewed and edited the output as needed and take full responsibility for the content of the publication.

